# Glove-related contact dermatitis: diagnostic value of a repeated application test

**DOI:** 10.1101/2023.07.17.23292561

**Authors:** Céline Lamouroux, Léa Bertolotti, Clio Coste, Pauline Pralong, Marine-Alexia Lefevre, Justine Pasteur, Aude Clément, Marie-Christine Ferrier Le Bouëdec, Barbara Charbotel, Jean-Baptiste Fassier, Marc Vocanson, Jean-François Nicolas, Florence Hacard, Audrey Nosbaum

## Abstract

**Background:** Gloves are frequently the cause of occupational hand dermatitis and are responsible for irritant (ICD) and allergic contact dermatitis (ACD). The diagnosis of ACD is based on patch tests to gloves (PTg), semi-open tests to gloves (SOg), and patch tests to glove allergens (PTal).

**Objective:** The aim was to define the contribution of a glove repeated application test (GRAT) in the diagnosis strategy of glove-related contact dermatitis.

**Methods:** This retrospective study included patients with hand eczema who wore gloves and who completed a GRAT in the context of allergological explorations. It consisted of applying a piece of the glove maintained by a non-adhesive strip during 10 consecutive nights on the same area of the anterior face of the forearm.

**Results:** Ten patients were diagnosed with ACD to gloves. This diagnosis was made on positive SOg, PTg, GRAT and PTal for 2 patients, positive PTal and GRAT for 3 patients, positive SOg and GRAT for 2 patients, positive GRAT alone for 2 patients, positive PTal alone for 1 patient. The GRAT had better sensitivity than the other tests.

**Conclusion:** GRAT seems useful in the diagnostic strategy of glove contact dermatitis and could be proposed when the diagnosis of glove-related contact dermatitis is suspected.

## Introduction

Hand dermatitis is a public health issue that affects 14.5 % of the general population [1]. It represents more than 80% of occupational dermatitis [2] and its cost is estimated to be 5 billion euros per year in Europe [3]. Among personal protective equipment (PPE) gloves are the most frequent cause of occupational hand dermatitis; they are responsible for 90% of PPE-related dermatoses [4] and are responsible for irritant (ICD) and allergic contact dermatitis (ACD) in 70 and 15% of cases, respectively [5]. They are involved in 40-70% of occupational allergies to rubber additives [6], but they contain many other allergens [7] and all types of gloves can cause allergic contact dermatitis, including natural and synthetic rubber (nitrile, butadiene, butyl, polyisoprene, neoprene, polychloroprene), vinyl, polyurethane, polyester, etc.

Patch tests (PT), as well as open and semi-open (SO) tests are the reference diagnostic tests for ACD [8]. They can be supplemented by use tests which, like the repeated open application test (ROAT), aim to reproduce an eczema lesion following the repeated application of the suspected allergenic compound on non-lesional skin. Use tests simulate the exposure pattern to allergens as close as possible to that in daily life and products can be tested as a whole. These are commonly used for cosmetics and topical drugs [9].

In the case of gloves, the diagnosis of ACD is based on patch tests to gloves (PTg), semi-open tests to gloves (SOg) and patch tests to glove allergens (PTal) present in the European baseline series (EBS) and the European rubber series (ERS) and additions [10,11]. However, there is, to our knowledge, no published data on the place of use tests in the exploration of contact dermatitis to gloves. The aim of the present study was therefore to define the contribution of a glove repeated application test (GRAT) in the diagnosis strategy of glove-related contact dermatitis.

## Methods

### Patients

This retrospective study included patients with hand eczema who wore gloves in personnel or occupational activities and who completed a GRAT in the context of allergological explorations between 2018 to 2022 in the allergology and dermatology departments of 4 university hospitals of the Auvergne-Rhône-Alpes region of southeast-central France (Lyon, Grenoble, Clermont-Ferrand, and Saint Etienne).

### Epicutaneous tests

The GRAT was explained to the patients during the initial consultation with a visual support detailing the protocol (Figure S1). For the GRAT, a piece of at least 3×3cm of new and clean glove was affixed to the anterior surface of the forearm free of any lesion, interior side towards the skin. The glove was held in contact with the skin by a strip of Nylex^®^-type fabric to maintain it without occlusion. The glove was applied in the evening at bedtime, maintained for a duration of 6 to 8 hours, and removed in the morning. The GRAT was repeated for 10 consecutive nights on the same area of skin. When possible, patients used a new piece of glove each night in case of disposable glove, otherwise the patients reused the same piece of glove. If an eczema lesion occurred during the GRAT, the patients were asked to contact the allergology and dermatology department to arrange consultation to confirm the positivity of the test. Positivity was defined by the presence of infiltrated erythema and papules or vesicles, covering at least 25% of the tested area, according to the positivity criteria of the repeated open application test (ROAT) proposed by Johansen et al. [12].

The semi-open glove tests (SOg) consisted of the application of a 3×3 cm piece of glove to the lesion-free area of the back of the patient, held in place by a sticking plaster for 2 days. The glove patch tests (PTg) consisted of the application of a glove punch (6mm) on the back of the patient (inner side in contact with the skin) under an aluminum (Finn chamber; Epitest Ltd,Oy, Tuusula, Finland) or plastic (IQ Ultra; Chemotechnique Diagnostics AB, Malmo, Sweden) cup for 2 days. Patch tests were also conducted using the extended European Baseline Series (EBS) and the European rubber series and additions (PTal) (Table S2). If patients used several types of gloves, all of them were tested. SOg, PTg, and PTal were left for 2 days and readings were made at Day 3 or Day 4 according to the European Society of Contact Dermatitis (ESCD) criteria. In order to assess delayed positive reactions, patients were instructed to report back on Day 7 if there was a suspected patch test reaction. Positive reactions were summarized from all reactions coded as +, ++, or +++, according to International Contact Dermatitis Research Group (ICDRG) definitions [8].

### Diagnosis of ACD

The diagnosis of ACD to gloves was made according to the following criteria: (i) at least one positive skin test to the patient’s gloves or to glove allergens; (ii) strong anamnestic clinical relevance; and (iii) disappearance of the contact dermatitis following the removal of the culprit glove and its replacement by another type of glove.

### Statistical analysis

Continuous data are reported as means (standard deviations) and categorical variables are reported as numbers (proportions). Sensitivity and specificity were calculated with their 95% confidence intervals and concordance of tests calculated using Cohen’s kappa coefficient. Analyses were conducted using the statistical software package SAS 9.4 (SAS Institute, Cary, NC, USA).

### Ethics

The participants received written information describing the study, its objectives, as well as the nature of the data collected, and were informed about their right to choose to participate or not. The study protocol was approved by the scientific and ethics committee of the Hospices Civils de Lyon (HCL, No. 22_5549 on 25/01/2022) and complied with the French data protection authority (*Commission nationale de l’informatique et des libertés*, CNIL) reference method MR004 and was registered under the number 22_5549 in the institutional CNIL register.

## Results

### 1. Patients and gloves

A total of 58 patients were included, 41 were female and the mean age was 37 years; 18 had a history of atopic dermatitis and 19 patients were healthcare workers. The mean time to consultation since the onset of eczema was 27 months (Table 1). A total of 84 gloves were tested. Forty-three were made of nitrile, 16 vinyl, 9 natural rubber, 4 neoprene, 2 polyisoprene, 3 a mixture of neoprene and polyisoprene, 1 polyester, and 1 polyurethane; for 5 gloves the composition was unknown (Table S1).

**Table 1:** Characteristics of the population.

|  | Total population<br>n = 58 |
| --- | --- |
| Sex, female, n (%) | 41 (70.7) |
| Mean age, years (SD) | 37 (12) |
| Atopy, n (%) |  |
| No | 39 (67.2) |
| Yes | 19 (32.8) |
| Atopic dermatitis, n (%) |  |
| No | 36 (62.0) |
| Yes | 18 (31.0) |
| NA | 4 (6.9) |
| Mean time to care, months (SD) | 27 (43) |
| Occupation, n (%) |  |
| Health care workers | 19 (32.8) |
| Hairdressers | 5 (8.6) |
| Automotive industry workers | 4 (6.9) |
| Metal industry workers | 3 (5.2) |
| Child daycare workers | 3 (5.2) |
| Dental personnel | 3 (5.2) |
| Chemists | 3 (5.2) |
| Cosmetologists | 2 (3.4) |
| Cleaners | 2 (3.4) |
| Cooks | 2 (3.4) |
| Clean room operators | 2 (3.4) |
| Construction workers | 2 (3.4) |
| Other | 8 (13.7) |

SD: standard deviation, NA: not available

### 2. Final etiological diagnosis

At the end of the allergological assessment, 10 patients were diagnosed as having ACD to gloves and 8 patients as having ACD to another product; 40 had a diagnosis other than ACD (Table S1).

### 3. ACD to gloves

Diagnosis of ACD to gloves was made on positive SOg, PTg, GRAT and PTal for 2 patients, positive PTal and GRAT for 3 patients, positive SOg and GRAT for 2 patients, positive GRAT alone for 2 patients, positive PTal alone for 1 patient (Table 2).

**Table 2:** Results of skin tests in the 10 patients with glove-related ACD.

| Patient | SOg | PTg | GRAT<br>(Day) | PTal | Glove type |
| --- | --- | --- | --- | --- | --- |
| 1 | 0 | 0 | + (D7) | 0 | neoprene +<br>polyisoprene |
|  | 0 | 0 | + (D10) |  | neoprene |
| 2 | + | NA | + (D2) | 0 | neoprene |
| 3 | + | + | + (NA) | Thiuram mix (+/-), TETD (+) | natural rubber |
| 4 | ++ | NA | + (D7) | 0 | neoprene |
| 5 | 0 | NA | + (NA) | Thiuram mix (+), TETD (+),<br>DPMTD (+), TMTM (++) | nitrile |
| 6 | 0 | 0 | + (D1) | Thiuram mix (++), TETD (++),<br>TMTM (++) | nitrile |
| 7 | 0 | 0 | + (NA) | Thiuram mix (+++), TMTD (++)<br>TETD (++) , DPMTD (++) ,<br>TMTM (++) , ZDMTC (+) | nitrile |
| 8 | 0 | 0 | + (D1) | 0 | nitrile |
| 9 | + | + | + (NA) | Thiuram mix (++), DPMTD (+), | natural rubber |
|  | 0 | 0 | + (NA) | TMTD (++) , TMTM (+++),<br>ZDMTC (+) | neoprene +<br>polyisoprene |
|  | 0 | 0 | + (NA) |  | neoprene |
| 10 | 0 | NA | 0 | Thiuram mix (++) | nitrile |

GRAT: Glove repeated application test, SOg: glove semi-open test, PTg: glove patch-test, PTal: glove allergen from European rubber series and additions and of the European standard baseline series, TETD: tetraethylthiuram disulfide, DPMTD: dipentamethylenethiuram disulfide, TMTM: tetramethylthiuram monosulfide, TMTD: tetramethylthiuram disulfite, ZDMTC: zinc dimethylthiocarbamate, NA: not available. Interpretation of patch tests and semi-open according to International Contact Dermatitis Research Group (ICDRG) definitions. Positivity of GRAT was defined by the presence of infiltrated erythema and papules or vesicles, covering at least 25% of the tested area

#### 3.1 Allergological tests with suspect gloves

##### 3.1.1 Glove semi open test (SOg)

There were 4 patients (#2, 3, 4, 9) who had positive a SOg test, all of which were associated with a positive GRAT and 2 (#3, 9) were associated with PTg and relevant PTal to thiurams. Positive SOg concerned 4 gloves: 2 in natural rubber and 2 in neoprene (Table 2). SOg had a <u>sensitivity of 40% (95%CI: 27-53) and a specificity of 100%.</u>

##### 3.1.2 Glove patch test (PTg)

Two PTg were positive, in patients who also had positive SOg, GRAT and relevant PTal to thiurams. Positive PTg concerned 2 gloves, in natural rubber (Table 3; patients #3 and 9). PTg had a <u>sensitivity of 33% (95%CI: 21-45) and a specificity of 100%.</u>

**Table 3:** Final diagnosis retained according to the results of skin tests Glove ACD (n=10) Other diagnosis (n=48)

|  | Glove ACD (n=10) | Other diagnosis (n=48) |
| --- | --- | --- |
| SOg POS | 4 | 0 |
| SOg NEG | 6 | 39 |
| SOg NA | 0 | 9 |
| PTg POS | 2 | 0 |
| PTg NEG | 4 | 30 |
| PTg NA | 4 | 18 |
| GRAT POS | 9 | 0 |
| GRAT NEG | 1 | 48 |
| PTal POS | 6 | 7 |
| PTal NEG | 4 | 37 |
| PTal NA | 0 | 4 |

SOg: glove semi-open test, PTg: glove patch-test, GRAT: Glove repeated application test, PTal: glove allergen from European rubber series and additions and of the European standard baseline series POS: positive, NEG: negative, NA: not available

##### 3.1.3 GRAT

Nine patients had a positive GRAT (Table 3). Two patients (#1 and 9) had positive GRAT on >1 glove. The time to GRAT positivity, known for 5 patients, ranged from 1 to 10 days, the mean interval was 4 days. Positive GRATs concerned 12 disposable gloves: 4 made of nitrile, 4 neoprene, 2 neoprene and polyisoprene, and 2 natural rubber.

For 2 patients (#1 and 8), the GRAT was the only positive test. Patient #1 was sensitized to 2 of the 3 types of gloves he used: 1 was neoprene, the second was neoprene and polyisoprene. The second patient (#8) used nitrile gloves. Patients #2 and #4 had positive SOg tests but PTal was negative while PTg was not performed. Patients #5, 6, and 7 were sensitized to several thiurams (PTal+) but SOg and PTg were negative. Patient #9 had GRAT+ for 3 gloves, the first was also SOg and PTg positive while the other 2 were negative with these tests; the patient was also sensitized to thiurams. For one patient (#10), the GRAT was negative while he had relevant sensitization to thiurams. There were 7/9 patients with a positive GRAT who had another relevant positive test (Sog, PTg, and/or PTal) to support the diagnosis of ACD (Table 2). The GRAT had a <u>sensitivity of 90% (95%CI: 82-98) and a specificity of 100%</u>.

#### 3.2. Patch tests with glove allergens (PTal)

Thirteen patients were sensitized to ≥1 glove allergens (Table S1; #3, 5, 6, 7, 9, 10, 19, 27, 28, 31, 39, 51, and 52). The allergens found were thiurams for 9 patients, including tetraethylthiuram disulfide (TETD) for 7 patients and tetramethylthiuram monosulfide (TMTM) for 6 patients. Patch test to dipentamethylenethiuram disulfide (DPMTD) was positive in 5 patients, tetramethylthiuram disulfide (TMTD) in 4 patients. Apart from thiurams, methylisothiazolinone was positive in 4 patients, and zinc dimethyldithiocarbamate (ZDMTC) and bisphenol A were positive in 2 patients. Patient #10 had positive patch test to thiuram mix but the detailed battery could not be performed. These sensitizations were considered relevant in 6 patients (#3, 5, 6, 7, 9, and 10; nitrile, neoprene, and natural rubber gloves; Table 3). Five of them (#3, 5, 6, 7, and 9) also had a positive GRAT, while in the other patient (#10) all tests performed with gloves (SOg and GRAT) were negative. In the other 7 patients (#19, 27, 28, 31, 39, 51, and 52) the final diagnosis ruled out ACD to gloves since the positivity for allergens did not correspond to the type of gloves, and all the skin tests using gloves were negative. Diagnoses were: 3 cases of irritant eczema, 2 cases of atopic eczema and 2 contact allergies to other occupational products (Table S1). The PTal had a <u>sensitivity of 60% (95%CI: 47-73) and a specificity of 84% (95%CI: 75-93).</u>

#### 3.3 Concordance between the GRAT and the other tests performed

As the results indicated that the GRAT had the greatest sensitivity for the diagnosis of glove ACD, the concordance between this test and the others performed was investigated (Table 4). Overall agreement was 90% between SOg tests and GRAT; κ was 0.58 (95%CI: 0.23-0.93). Three patients had positive GRAT with positive SOg as well, and for 6 patients who had a positive GRAT the SOg was negative.

**Table 4:** Comparison of the results obtained by GRAT to that obtained by the other glove tests.

|  | GRAT POS* | GRAT NEG | Total |
| --- | --- | --- | --- |
| SOg POS* | 4 | 0 | 4 |
| SOg NEG | 5 | 40 | 45 |
| SOg NA | 0 | 9 | 9 |
| PTg POS* | 2 | 0 | 2 |
| PTg NEG | 4 | 30 | 34 |
| PTg NA | 3 | 19 | 22 |
| PTal POS* | 5 | 8 | 13 |
| PTal NEG | 4 | 37 | 41 |
| PTal NA | 0 | 4 | 4 |

\* At least one positive test if several tested gloves per patient.
GRAT: glove repeated application test, SOg: glove semi-open test, PTg: glove patch-test, PTal: glove allergen from European rubber series and additions and of the European standard baseline series, POS: positive, NEG: negative, NA: not available

Six patients with positive GRAT underwent a PTg; one had a positive PTg (patient #9). The overall agreement between GRAT and PTg was 89%; κ was 0.45 (95%CI: -0.06 to 0.96).

For 5 patients, PTal and GRAT were positive, for the other 4 patients with positive GRAT, PTal was negative. PTal was positive for 6 patients with negative GRAT; 1 was considered clinically relevant (Table 2; patient #2). The overall agreement between GRAT and PTal was 81%; κ was 0.33 (95%CI: 0-0.66).

### 4 ACD to other compounds

Eight patients were allergic to products other than gloves or glove allergens: acrylates known to be contained in nail varnish for a nail esthetician (patient #16), chromium and cobalt known to be contained in cement for a builder (#38), bisphenol A for a textile impregnation operator (#39), tensioactives (lauryl glucoside, decyl glucoside) contained in shampoos used by a hairdresser (#40), benzisothiazolinone contained in a lubricant used by a dental technician (#41), paraphenylenediamine (PPD) in a hair dye used by a hairdresser (#47), cutting oils used by machinist (#52), and another product than glove, which was not specified by the reporting physician, for a nurse (#46; Table S1).

### 5 Other diagnoses

The diagnoses retained in the other 40 patients with hand eczema were: ICD for 16 patients, AD for 14 patients, mixed eczema (AD + ICD) for 9. For one patient (#37), who had an isolated and brief episode of eczema, the etiology was not identified (Table S1).

## Discussion

The results of the present study show that CD to gloves represents a significant proportion of the cases of hand eczema in glove wearers. Of note, ACD affected a third of patients, which is in keeping with that reported recently in a large study that found that for 28% of patients hand eczema had an allergic etiology [13]; furthermore, the GRAT was the most frequently positive glove test, followed by SOg, whereas glove PTg was rarely positive.

The PTal allowed the characterization of the allergen involved in the eczema when positive. The most frequently found allergens were glove allergens; thiurams, methylisothiazolinone, zinc dimethyldithiocarbamate, and bisphenol A. This concerned nearly a quarter of patients, which is related to the inclusion of patients who used gloves, but it should be noted that such individuals are prone to be allergic to other rubber accelerators such as other dithiocarbamates, mercaptobenzothiazole as well as its derivatives, diphenylguanidine and thioureas, but also that vinyl gloves expose users to bisphenol A, formaldehyde, benzisothiazolinone, isobutyrates, polyadipates, dyes, and phthalates, while leather gloves expose users to chromium and metal gloves to nickel [6]. However, skin tests using personal gloves also contributed to the diagnosis of ACD, in keeping with a previous study [14]. <u>One major finding of the present study is the excellent sensitivity of the GRAT that was better than the other tests using patients’ gloves</u> (i.e. GRAT > SOg > PTg); the GRAT detected 9/10 glove ACD while the SOg diagnosed only 4, and PTg 2. Several hypotheses could explain this intriguing observation. First, the larger area of the test material in the GRAT and SOg (3×3 cm = 9 cm²) compared to PTg (6 mm diameter = 50 mm²) may lead to increase the release of glove allergens and therefore enhanced reactivity. Likewise, in the study reported by Basch et al. comparing the PT results according to the size of the Finn chamber (large versus small) concluded that large test chambers may be useful for detection of weak sensitizations to particular contact allergens [15]; this was later confirmed in a subsequent study reported by Geffeler et al. who found that a larger contact surface area between a hapten and the skin led to an increase in PT reactivity [16]. Second, the superiority of the GRAT could be explained by the anatomical area tested; the skin of the forearm (GRAT) is thinner than that of the back (PTg and SOg) and would therefore allow more allergens to penetrate. Third, repeating the test daily with a new piece of glove for 10 nights (GRAT) rather than applying a single piece left for 2 days (SOg, PTg), would allow optimal delivery of the allergen. Along the same line, the GRAT is the adaptation to gloves of the ROAT, reported by Hannuksela and Salo in 1986 to clarify doubtful or inconsistent results of PT [17]. Like the ROAT, <u>the GRAT has the advantage of being a simple and non-invasive test that mimics the usage situation as much as possible</u> to increase the ability of detecting ACD. Concerning the GRAT protocol, we chose a duration of 10 nights because it seemed to be a good compromise between the sensitivity of the test and the adherence of the patients; the Dermato-Allergology Group of the French Society of Dermatology had shown in a recently published study that 98% of positive ROAT occurred within 10 nights of application of the product [18]. Although we minimized the occlusion properties of the test by using a non-adhesive strip and a shorter application time, repetitive application of a new piece of glove for 10 nights could induce innate skin inflammation (skin irritation) in predisposed patients. However, <u>there are several reasons to think that a positive GRAT reflects contact sensitization</u>: first, a recent study reported that the gloves have only a very low cutaneous irritant potential and the isolated occlusion is qualified as low irritant [19] when other irritating agents or factors (washing, soaps, creams, etc.) involved in glove eczema are eliminated; second, the time to GRAT positivity in the present study was short (mean 4 days), while no signs of skin inflammation were noted after 10 days in the remaining 49 patients, suggesting that the GRAT procedure is not irritating; third, replacing the incriminated gloves with another type of glove resulted in rapid healing of the lesions in all patients with a positive GRAT; fourth, 7/9 patients with positive GRAT had another relevant positive test (SOg, PTg, and/or PTal) to support the diagnosis of ACD. For the other 2 patients (#1 and #8), the GRAT was the only positive test and its clinical presentation was strongly suggestive of sensitization with the presence of several papules and vesicles. In order to confirm the sensitization hypothesis, we carried out, in patient #1, a biopsy of the active eczematous lesions of the GRAT for molecular analysis of the cutaneous inflammation making it possible to differentiate allergic eczema from irritant eczema [20]. This confirmed the diagnosis of GRAT-induced contact allergy by showing typical features of ACD, i.e. skin infiltration and activation of cytotoxic CD8+ T cells (*data not shown*). Unfortunately, the other patient (#8) did not undergo this molecular analysis. However, replacing the offending gloves with another type of glove resulted in a rapid improvement in his hand eczema. Thus, the GRAT was essential to the diagnosis of glove contact dermatitis in these 2 patients.

### Limitations of the study

Although the GRAT was performed by most of the patients to whom it was proposed, there is no control over whether they followed precisely the protocol at home. Negative results should take this into account; for example, patient #3 with glove ACD revealed by sensitization to thiuram mix had a negative GRAT. He had difficulty understanding French language and it is suspected that he did not perform the test correctly. A good understanding of the test and the patient’s cooperation seem to be the main limitations of this test. A clear explanation, presenting the purpose of the test, with the use of visual aids seems necessary. Although the procedure was summarized graphically in an information sheet (Figure S1) this could be improved with a video explaining the procedure. More generally, the sample size does not allow us to perform statistical analyses sufficiently extensive to propose robust statistical outliers for the GRAT and to assert that it differentiates allergy to irritation.

## Conclusion

GRAT is a use test that seems useful in the diagnostic strategy of glove contact dermatitis. Its non-invasive character and its easy realization allow it to be proposed when the diagnosis of glove-related contact dermatitis is suspected.

**Figure 1:**
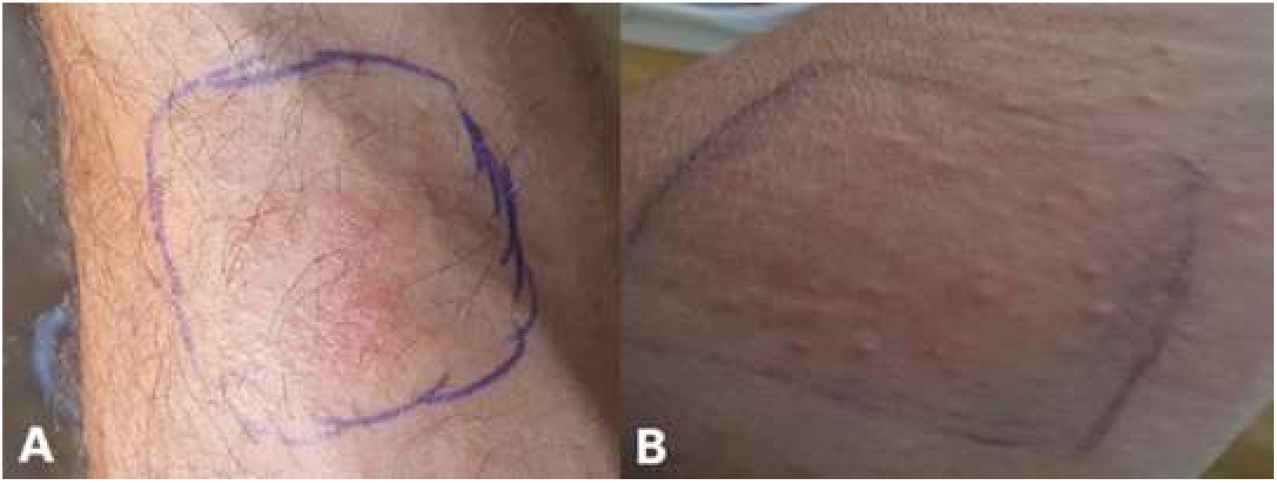
Example of positive GRAT positive results. AA A) Patient #6; GRAT with a nitrile glove, on day 2. B) Patient #2; GRAT with a neoprene glove, on day 3.

## Data Availability

All data produced in the present study are available upon reasonable request to the authors

## Acknowledgements

The authors thank Dominique Tennstedt (Bruxelles) and Marie-Noëlle Crépy (Créteil) for their critical reading of the manuscript.

## Supplemental material

Table S1: Details of patient characteristics

Table S2: Allergens used in patch tests

Figure S1: Visual GRAT protocol for patients

