## Supplemental Table S1 for "Glove-related contact dermatitis: diagnostic value of a repeated application test"

| N | Sex | Age range | Atopy | AD | Job | Time to consultation (months) | Glove type | PT EBS | PTal | PTg | SOg | GRAT (day) | Diagnosis |
|---|---|---|---|---|---|---|---|---|---|---|---|---|---|
| **1** | M | 56-60 | NO | NO | Nurse | 3 | natural rubber | 0 | 0 | 0 | 0 | NEG | ACD |
| | | | | | | | neoprene + polyisoprene | | | 0 | 0 | POS (D7) | |
| | | | | | | | neoprene | | | 0 | 0 | POS (D10) | |
| **2** | F | 46-50 | NA | YES | Sawing technician | 14 | neoprene | Nickel (+) | 0 | NA | (+) | POS (D2) | ACD |
| **3** | F | 26-30 | NO | YES | Dental assistant | NA | natural rubber | 0 | Thiuram mix (+/-), TETD (+) | (+) | (+) | POS | ACD |
| | | | | | | | nitrile | | | 0 | 0 | NEG | |
| **4** | M | 56-60 | YES | YES | Metal worker | 168 | neoprene noir | Potassium dichromate (++) MIT (+) | 0 | NA | (++) | POS (D7) | ACD |
| **5** | F | 41-45 | NO | NO | Laboratory technician | 12 | nitrile | 0 | Thiuram mix (+) TETD (+) DPMTD (+) TMTM (++) | NA | 0 | POS | ACD |
| **6** | M | 36-40 | NO | NO | Auto mechanic | 7 | nitrile | MIT + MCIT (++), MIT (+++), OIT (+) | Thiuram mix (++), TETD (++), TMTM (++) | 0 | 0 | POS (D1) | ACD |
| **7** | F | 31-35 | NO | NO | Assistant laboratory engineer | 48 | nitrile | PPD (+) | Thiuram mix (+++), TMTD (++), TETD (++), DPMTD (++), TMTM (++), ZDMTC (+) | 0 | 0 | POS | ACD |
| | | | | | | | nitrile | | | 0 | 0 | POS | |
| | | | | | | | nitrile accelerator free | | | 0 | 0 | NEG | |
| **8** | F | 31-35 | NO | NO | Process technician in clean room | 8 | nitrile | 0 | 0 | 0 | 0 | POS (D1) | ACD |
| **9** | F | 21-25 | NO | NO | Surgery resident | 12 | natural rubber | 0 | Thiuram mix (++), DPMTD (+), TMTD (++), TMTM (+++), ZDMTC (+) | (+) | (+) | POS | ACD |
| | | | | | | | neoprene + polyisoprene | | | 0 | 0 | POS | |
| | | | | | | | neoprene | | | 0 | 0 | POS | |
| | | | | | | | neoprene accelerator free | | | 0 | 0 | NEG | |

| | | | | | | | | | | | | | |
|---|---|---|---|---|---|---|---|---|---|---|---|---|---|
| **10** | M | 56-60 | NO | NO | Hospital cook | 6 | nitrile | 0 | Thiuram mix (++; D7) | NA | 0 | NEG | ACD |
| | | | | | | | nitrile | | | NA | 0 | NEG | |
| **11** | F | 36-40 | NO | NO | Chemical engineer | 10 | nitrile | 0 | NA | NA | 0 | NEG | AD+ICD |
| | | | | | | | nitrile | | | NA | 0 | NEG | |
| **12** | M | 36-40 | NO | NO | Forklift operator | 21 | polyester | 0 | 0 | NA | NA | NEG | ICD |
| **13** | F | 21-25 | YES | YES | Cooking | 36 | natural rubber | 0 | 0 | 0 | 0 | NEG | AD |
| **14** | F | 31-35 | NO | NO | Nurse | 12 | nitrile | Nickel (+++) | 0 | NA | 0 | NEG | AD |
| | | | | | | | vinyl | | | NA | 0 | NEG | |
| | | | | | | | natural rubber | | | NA | 0 | NEG | |
| **15** | F | 36-40 | YES | YES | Gynecologist-obstetrician | 180 | nitrile | 0 | 0 | 0 | 0 | NEG | AD |
| | | | | | | | powder nitrile | | | 0 | 0 | NEG | |
| | | | | | | | vinyl | | | 0 | 0 | NEG | |
| | | | | | | | neoprene + polyisoprene | | | 0 | 0 | NEG | |
| | | | | | | | natural rubber | | | 0 | 0 | NEG | |
| | | | | | | | polyisoprene | | | 0 | 0 | NEG | |
| **16** | F | 31-35 | NO | NO | Nail technician apprentice | 6 | nitrile | Nickel (++) | 0 | NA | 0 | NEG | ACD |
| **17** | M | 46-50 | NO | NO | Rectifier technician | 3 | nitrile | 0 | 0 | 0 | NA | NEG | ICD |
| **18** | F | 31-35 | NO | NO | Childcare assistant | 12 | vinyl | 0 | 0 | NA | 0 | NEG | ICD |
| **19** | F | 31-25 | NO | YES | Care assistant | 36 | vinyl | 0 | Thiuram mix (+), TMTD (+++; D3) MDA (+; D3), TETD (++; D3) DPMTD (+; D3), CPPD (+; D4), TMTM (+++; D3) | NA | 0 | NEG | ICD |
| **20** | M | 46-50 | NO | NO | Metal rectifier | 22 | nitrile | 0 | 0 | NA | 0 | NEG | AD+ICD |
| **21** | F | 36-40 | NO | NO | Nurse | 72 | nitrile | 0 | 0 | NA | 0 | NEG | AD+ICD |
| **22** | F | 31-35 | NO | NO | Nurse | 3 | nitrile | 0 | 0 | NA | 0 | NEG | ICD |
| **23** | F | 36-30 | NO | NO | Cleaning worker | 12 | nitrile | 0 | 0 | NA | 0 | NEG | ICD |
| | | | | | | | nitrile | | | NA | 0 | NEG | |

| | | | | | | | | | | | | | |
|---|---|---|---|---|---|---|---|---|---|---|---|---|---|
| **24** | F | 41-45 | YES | YES | Care assistant | 120 | nitrile<br>nitrile | 0 | NA | NA<br>NA | NA<br>NA | NEG<br>NEG | AD |
| **25** | F | 45-50 | NO | NO | Care assistant | 24 | vinyl<br>nitrile | 0 | 0 | NA<br>NA | NA<br>NA | NEG<br>NEG | AD |
| **26** | F | 21-25 | NO | NO | Nurse | 9 | vinyl | 0 | NA | NA | NA | NEG | ICD |
| **27** | M | 61-65 | YES | YES | Retired | 2 | polyurethane + nylon | Peru balsam (+)<br>Cocamide DEA (+)<br>Linalol hydroperoxyde (+++) | Formaldehyde (++) | NA | 0 | NEG | AD |
| **28** | F | 21-25 | YES | YES | Care assistant | 7 | vinyl | Quaternium 15 (+; D7)<br>Textile dye mix (+; D7) | TETD (++; D7),<br>DNPD (+; D7)<br>DPT (+; D7),<br>BPA (+; D7) | NA | NA | NEG | AD |
| **29** | F | 26-30 | YES | YES | Childcare assistant | 3 | vinyl<br>nitrile | 0 | 0 | NA<br>NA | 0<br>0 | NEG<br>NEG | AD |
| **30** | F | 41-45 | NO | NO | Nurse | 36 | nitrile | Nickel (++)<br>MIT+MCT (++)<br>Textile dye mix (+)<br>MIT (++) | 0 | NA | 0 | NEG | AD |
| **31** | F | 46-50 | NO | NO | Dental assistant | 9 | vinyl | Nickel (++)<br>MDBGN (++)<br>Textile dye mix (+) | Thiuram mix (+),<br>TMTD (+), TETD (++),<br>DPMTD (++), ZDETC (+<br>TMTM (+) | 0 | 0 | NEG | ICD |
| **32** | F | 21-25 | NO | NO | Jeweler | 3 | nitrile | 0 | 0 | 0 | 0 | NEG | ICD |
| **33** | F | 21-25 | YES | YES | Fast food employee | 3 | vinyl | 0 | 0 | 0 | 0 | NEG | AD |
| **34** | F | 56-60 | NO | NO | Technical agent | 6 | MAPA | Nickel (+++) | 0 | 0 | 0 | NEG | AD+ICD |
| **35** | F | 51-55 | YES | YES | Financial inspector | 3 | vinyl | 0 | 0 | 0 | 0 | NEG | AD |
| **36** | M | 21-25 | NO | NO | Clean room technician | 7 | nitrile | 0 | 0 | 0 | 0 | NEG | AD |
| **37** | M | 21-25 | NO | NO | Car painter and body repairer | 2 | nitrile noir | 0 | 0 | 0 | 0 | NEG | NA |
| **38** | M | NA | NO | NA | Mason | 8 | vinyl<br>textile + | Potassium dichromate (++), | 0 | NA<br>NA | NA<br>NA | NEG<br>NEG | ACD |

| | | | | | | | | | | | | | |
|---|---|---|---|---|---|---|---|---|---|---|---|---|---|
| | | | | | | | rubber (unspecified) | cobalt (+) | | | | | |
| **39** | M | 46-50 | NO | NO | Operator in textile impregnation | 4 | nitrile | Budesonide (+) | BPA (++) | 0 | 0 | NEG | ACD |
| **40** | F | 16-20 | YES | YES | Apprentice hairdresser | 5 | natural rubber | Lauryl glucoside (+), decyl glucoside (+) | 0 | 0 | 0 | NEG | ACD |
| **41** | F | 36-40 | NO | NO | Dental technician | 4 | vinyl | MIT (++), MDBGN (++ | 0 | 0 | 0 | NEG | ACD |
| **42** | M | 41-45 | NO | NO | Chimney installer | 48 | textile | 0 | 0 | 0 | 0 | NEG | AD |
| **43** | F | 26-30 | YES | YES | Hairdresser | 156 | nitrile | 0 | 0 | 0 | 0 | NEG | AD+ICD |
| **44** | F | 31-35 | YES | YES | Hairdresser | 11 | nitrile black | 0 | 0 | 0 | 0 | NEG | ICD |
| **45** | F | 16-20 | YES | YES | Hairdresser | NA | NA | 0 | 0 | 0 | 0 | NEG | AD+ICD |
| **46** | F | 21-25 | YES | YES | Nurse | NA | nitrile | 0 | 0 | 0 | 0 | NEG | AD+ACD |
| | | | | | | | nitrile | | | 0 | 0 | NEG | |
| **47** | F | 46-50 | YES | NO | Hairdresser | NA | nitrile | PPD | 0 | 0 | 0 | NEG | ACD |
| **48** | M | 36-40 | NO | NO | Mechanic | NA | nitrile | 0 | 0 | 0 | 0 | NEG | ICD |
| **49** | M | 36-40 | NO | NO | Waste sorting center agent | NA | nitrile | Nickel (++) | 0 | 0 | 0 | NEG | ICD |
| **50** | F | 21-25 | YES | NO | Nurse in training | 12 | nitrile | 0 | 0 | 0 | 0 | NEG | ICD |
| **51** | F | 36-40 | NO | NO | Anatomical Pathology Laboratory Preparer | 12 | nitrile | Linalol hydroperoxyde (+) | Carba mix (++) | 0 | 0 | NEG | ICD |
| **52** | M | 56-60 | NO | NO | Mechanic (machining) | 60 | handling (unspecified) | Limonene hydroperoxyde (+/-) Workshop cutting fluids | Carba mix (+) | 0 | 0 | NEG | ACD |
| **53** | F | 41-45 | NO | NO | Childcare assistant | 8 | MAPA | 0 | 0 | 0 | 0 | NEG | ICD |
| | | | | | | | nitrile | | | 0 | 0 | NEG | |
| **54** | F | 46-50 | NO | NO | Hospital Service Officer | 8 | vinyl | Nickel (++) | 0 | 0 | 0 | NEG | ICD |
| | | | | | | | nitrile | | | 0 | 0 | NEG | |
| **55** | F | 21-25 | YES | YES | Care assistant | 10 | vinyl | 0 | 0 | 0 | 0 | NEG | AD+ICD |
| | | | | | | | nitrile | | | 0 | 0 | NEG | |
| **56** | F | 36-40 | NO | NA | Esthetician | 2 | nitrile | Cobalt | NA | 0 | NA | NEG | AD+ICD |

| | | | | | | | | | | | | |
|---|---|---|---|---|---|---|---|---|---|---|---|---|
| **57** | M | 21-25 | YES | NA | Site foreman | 12 | leather, plastic, tissu | 0 | 0 | 0 | NA | NEG | AD |
| **58** | F | 56-60 | NO | NA | Cleaning worker | 120 | vinyl | 0 | 0 | 0 | 0 | NEG | AD+ICD |

Table S1: Details of patients' characteristics. GRAT: glove repeated application test, SOg: glove semi-open test, PTg: glove patch-test, PT EBS : Patch test from European Baseline series, PTal: glove allergen patch test from European rubber series and additions, TETD: tetraethylthiuram disulfide, MIT: methylisothiazolinone, OIT: octylisothiazolinone, DPMTD: dipentamethylenethiuram disulfide, TMTM: tetramethylthiuram monosulfide, MCIT: methylchloroisothiazolinone, TMTD: tetramethylthiuram disulfite, ZDMTC: zinc dimethylthiocarbamate, PPD: p-phenylenediamine, MDA: diaminodiphenylmethane, CPPD: cyclohexyl-N-phenylphenylenediamine, BPA: bisphenol A, DNPD: di-naphtylphenylenediamine , DPT: diphenylthiourea, ZDETC: zinc diethylthiocarbamate, MDBGN: methyldibromoglutaronitrile,  NA: not available. Interpretation of patch tests and semi-open according to International Contact Dermatitis Research Group (ICDRG) definitions. Positivity of GRAT was defined by the presence of infiltrated erythema and papules or vesicles, covering at least 25% of the tested area. POS: positive, NEG: negative. ACD: allergic contact dermatitis, ICD: irritant contact dermatitis, AD: atopic dermatitis
