## Supplemental Table S2 for "Glove-related contact dermatitis: diagnostic value of a repeated application test"

Table S2: Allergens used in patch tests

| European baseline series | European rubber series and additives |
|---|---|
| Potassium dichromate 0.5% | Tetramethylthiuram disulfite 1% |
| Neomycin sulfate 20% | Diaminodiphenylmethane 0.5% |
| Thiuram mix 1% | Tetramethylthiuram disulfide 1% |
| P-Phenylenediamine 1% | Cyclohexyl-Benzothiazolsulfenamide 1% |
| Cobalt chloride 1% | Dipentamethylenethiuram disulfite 1% |
| Methyldibromo glutatronitrile 0.5% | Dibenzothiazyl Disulfide 1% |
| Formaldehyde 2% | Morphonylmercaptobenzothiazol 1% |
| Colophonium 20% | Zinc dibuthyldithiocarbamate 1% |
| Benzocaine 5% | Cyclohexyl-phenylphenylenediamine 1% |
| Peru balsam 25% | Diphenylguanidine 1% |
| N-Isopropyl-phenyl-paraphenylenediamine 0.1% | Carba mix (n,n-diphenylguanidine, zinc diethyldithiocarbamate, zinc dibutyldithiocarbamate) 3% (in one center) |
| Lanolin alcohol 30% | Di-naphtyl-phenylenediamine 1% |
| Mercapto mix 2% | Zinc diethyldithiocarbamate 1% |
| Bisphenol A 1% | Methenamine (Hexamehylenetetramine) 2% |
| Paraben mix 16% | Tetramethylthiuram monosulfide 1% |
| Tert –butylphenolformaldehyde resin 1% | Diphenylthiourea 1% |
| Fragrance mix 1 8% | Zinc Dimethyldithiocarbamate 1% |
| Clioquinol 5% | Diethylthiourea 1% |
| Quaternium 15 1% | Ethylenediamine dihydrochloride 1% |
| Nickel sulfate 5% | Dibutylthiourea 1% |
| Methylchlorisothiazolinone + Methylisothiazolinone 0.02% | Phenyl-naphtylamine 1% |
| Mercaptobenzothiazole 2% | Diphenyl-p-phenylenediamine 1% |
| Methoxy–pentyl–benzoquinone 0.01% | Cyclohexylthiophtalimide 1% |
| Budesonide 0.01% | Ter-Butylcathecol 0.25% |
| Tixocortol 21 pivalate 0.1% | Benzisothiazolinone 0.1% |
| Cocamidopropyl betaine 1% | Cetylpyridinium chloride 0.1% |
| Dimethylaminopropylamine 1% | Tricresylphosphate 5% |
| Hydroxyisohexyl-cyclohexene carboxaldehyde 5% | Dioctylphtalate 2% |
| Cocamide DEA 0.5% | Triphenylphosphate 5% |
| Armerchol L 101 50% | Bisphenol A 1% |
| Fragrance Mix 2 14% | Dibutylphtalate 5% |
| Sesquiterpene lactone Mix 0.1% | |
| Hydroperoxyde de Linalol 1% | |
| Benzyl alcohol 10% | |
| Limonene hydroperoxyde 0.3% | |
| Textile dye Mix 6.6% | |
| Lauryl polyglucose 3% | |
| Methylisothiazolinone 0.2% | |
| Octylisothiazolinone 0.1% | |
| Benzalkonium chloride 0.1% | |
