## Supplementary figures and images for "Glove-related contact dermatitis: diagnostic value of a repeated application test"

### Supplemental Figure S1

Figure S1: Visual GRAT protocol for patients

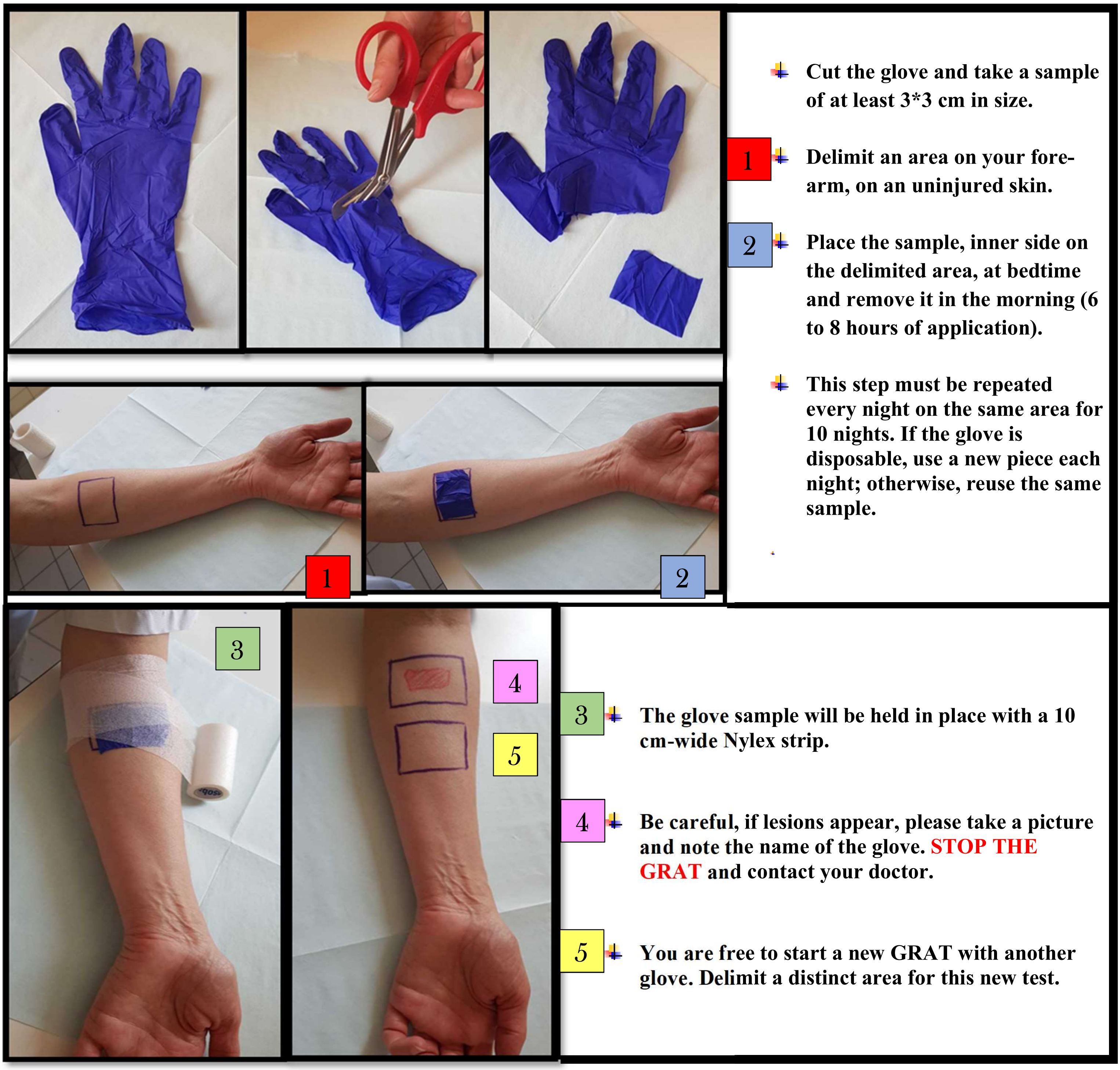
